# Bio-efficacy of field aged novel class of long-lasting insecticidal nets, against pyrethroid-resistant malaria vectors in Tanzania: A series of experimental hut trials

**DOI:** 10.1101/2023.10.21.23297289

**Authors:** Jackline L. Martin, Louisa A. Messenger, Mark Rowland, Franklin W Mosha, Edmund Bernard, Monica Kisamo, Shaban Limbe, Patric Hape, Charles Thickstun, Crene Steven, Oliva Moshi, Boniface Shirima, Nancy S. Matowo, Jacklin F Mosha, Dominic P Dee, Thomas S Churcher, Manisha A. Kulkarni, Alphaxard Manjurano, Natacha Protopopoff

## Abstract

New classes of long-lasting insecticidal nets (LLINs) incorporating two insecticides, or an insecticide and a synergist, are recommended by the World Health Organization (WHO) to prevent malaria transmitted by mosquito vectors resistant to pyrethroid and other common insecticide classes. This study was nested in a large-scale cluster-randomized controlled trial conducted in Tanzania. A series of experimental hut trials (EHTs) aimed to evaluate the bio-efficacy of trial LLINs on the mosquito indicators most pertinent to malaria transmission over 3 years of use in the community. The aim was to evaluate nets subjected to a broader range of household factors than WHO standardized washing.

The following field collected LLINs were assessed: 1/Olyset^TM^ Plus (combining piperonyl butoxide synergist and permethrin), 2/Interceptor^®^ G2 (chlorfenapyr and alpha-cypermethrin), 3/Royal Guard^®^ (pyriproxyfen and alpha-cypermethrin), 4/Interceptor^®^ (alpha-cypermethrin only), 5/a new Interceptor^®^, and 6/an untreated net. Thirty nets of each type were withdrawn from the community at 12, 24 and 36 months after distribution and used for the EHTs. Pre-specified outcomes were 72-hour mortality for Interceptor^®^ G2, 24-hour mortality for Olyset^TM^ Plus, and fertility based on egg development stage for Royal Guard^®^.

Overall; Interceptor^®^ G2 LLINs induced higher 72-hour mortality compared to standard LLINs of the same age up to 12 months (44% vs 21%), OR: 3.5, 95% CI: 1.9 – 6.6, p-value < 0.001 and 24-hour mortality was only significantly higher in Olyset^TM^ Plus when new (OR: 13.6, 95%CI: 4.4 – 41.3, p-value < 0.001) compared to standard LLINs but not at 12 months (17% vs 13%; OR: 2.1, 95% CI: 1.0 – 4.3; p-value = 0.112). A small non-significant effect of pyriproxyfen on *Anopheles* fertility was observed for Royal Guard^®^ up to 12 months (75% vs 98%, OR: 1.1, 95% CI: 0.0 – 24.9, p-value = 0.951). There was no evidence of a difference in the main outcomes for any of the new class of LLINs at 24 and 36 months compared to standard LLINs.

Interceptor^®^ G2 LLINs showed superior bio-efficacy compared to standard LLINs for only up to 12 months and the effect of Olyset^TM^ Plus was observed when new for all species and 12 months for *An. gambiae* s.l. only. The pyriproxyfen component of Royal Guard^®^ had a short and limited effect on fertility

## Introduction

Malaria remains a considerable global problem, with an estimated 241 million cases in 2020, resulting in 627,000 deaths. The African region of the World Health Organization (WHO) accounted for 96% of all malaria cases and related deaths (1). To mitigate burden of malaria, 2.3 billion insecticide-treated nets (LLINs) have been distributed worldwide between 2004 and 2022, with 86% delivered to sub-Saharan Africa (1). In 2020, malaria endemic countries received an estimated 229 million LLINs, with 19.4 million of them being pyrethroid-piperonyl butoxide (PBO)-treated nets (1). LLINs and other vector control interventions are predicted to have averted 1.7 billion malaria cases and 10.6 million malaria deaths between 2000 and 2020 (1). Coincident with the scaling up of LLINs and other insecticidal tools across sub-Saharan Africa, mosquito vector populations have developed resistance to pyrethroids, which until recently were the only insecticide class recommended for use in LLINs. Several studies have demonstrated that LLINs are becoming less effective at killing mosquitoes in areas of high pyrethroid resistance (2–4). To tackle the spread of resistance, new malaria vector control tools were needed. The first new class of LLINs developed to control resistant mosquitoes was a combination net containing a pyrethroid insecticide and the synergist PBO (5). In experimental field trials, these LLINs induced significant mortality in vector species (6). In Burkina Faso, Olyset^TM^ Plus outperformed pyrethroid-only LLIN (7). This class of LLINs was initially evaluated in a cluster-randomized control trial (cRCT) in Tanzania and Uganda. In Tanzania, the study compared Olyset^TM^ Plus to Olyset^TM^ net and found a 44% reduction malaria prevalence in the pyrethroid-PBO LLIN arm (Olyset^TM^ Plus) compared to the standard pyrethroid LLIN arm (Olyset^TM^ net) arm after one year, a 33% reduction after two years (8). In the Ugandan study, conducted over 18 months, the pyrethroid-PBO LLIN (PermaNet^®^ 3.0, Olyset^TM^ Plus, PermaNet^®^ 2.0 and Olyset ^TM^ net) showed a 14% reduction in prevalence. The former results led the WHO to recognize the public health value of pyrethroid-PBO LLINs and provide a conditional recommendation as a new class of vector control product (9).

Since then, two additional dual-active ingredient (AI) LLINs (Royal Guard^®^ and Interceptor^®^ G2) have undergone evaluation in WHO Phase I and II trials, showing significant promise in comparison to standard LLINs when combating pyrethroid-resistant vectors. In Phase I studies, Royal Guard^®^ (containing the pyrethroid alpha-cypermethrin, and the juvenile growth hormone inhibitor pyriproxyfen) met the WHO criteria with 95% knockdown and more than 80% mortality for up to 20 washes when the susceptible *Anopheles gambiae* s.s. (kisumu strain) were exposed in cone assays (10). It also met the WHO criteria in tunnel tests, with mortality exceeding 80% after 20 washes, in contrast to a pyrethroid-only net. In a Phase II experimental hut trial conducted against wild, pyrethroid resistant *An gambiae* s.l., Royal Guard^®^ demonstrated an 83% reduction in oviposition and a 95% reduction in offspring/hatching before washing. These values declined to 25% and 50%, respectively, after 20 washes. Interceptor^®^ G2 (containing alpha-cypermethrin and the pyrrole chlorfenapyr) was able to induce 71% mortality against free flying *An. gambiae* s.l. in an experimental hut trial compared to an alpha-cypermethrin-only net (20% mortality) (11).

In a large-scale cRCT conducted in Tanzania, Royal Guard^®^, Interceptor^®^ G2 and Olyset^TM^ Plus LLINs were evaluated against standard Interceptor^®^ nets in the context of resistant malaria vectors. The results revealed 55% lower odds of malaria infection in children aged 6 to 14 years after two years of net use in the Interceptor^®^ G2. Additionally, the entomological inoculation rate (EIR) was also significantly lower (by 85%) in the Interceptor^®^ G2 arm compared to the standard pyrethroid-only arm (12). In the same trial, Olyset^TM^ Plus showed a shorter protective effect of 12 months compared to a previous cRCT conducted in a different part of Tanzania, where pyrethroid-PBO LLINs remained effective for 24 months (8, 12).

A second cRCT conducted in Benin showed similar results, with Interceptor^®^ G2 reducing malaria incidence by 44% compared to pyrethroid-only LLINs, while Royal Guard^®^ did not significantly reduce malaria outcomes (13). Among the new dual-AI LLINs evaluated in different settings, Interceptor^®^ G2 received strong recommendation to be deployed for malaria prevention to children and adults. In contrast, Royal Guard^®^ received a conditional recommendation for use as an alternative to standard pyrethroid-only nets.

During prospective WHO Phase II evaluations of LLINs efficacy, it is expected that each net should retain insecticidal activity for at least 20 standardized washes, with respect to vector knock-down, mortality, and blood feeding inhibition (14). However, studies done using dual-AI LLINs in experimental huts reported improved personal protection. It is important to note that all these studies were caried out using unwashed nets, with the nets washed 20 times as a proxy for a 3-year net used in the community. It is worth considering that insecticide retention on nets in community may be lower than in LLINs washed 20 times, as community washing process can be more intense and the nets more subject to friction during general use (17). In addition, hole size in aged nets in community may differ from the 4 x 4 cm standardized holes created in washed nets. Data from experimental hut trials conducted with ‘real-life’ field nets can help explain why Royal Guard^®^ LLINs had a more moderate effect on mosquito density and transmission in community trials, and why Olyset^TM^ Plus LLINs did not last for more than 24 months in recent trials (12, 15).

Through a series of experimental hut trials, we aimed to evaluate the bio-efficacy of dual-AI LLINs (Olyset^TM^ Plus, Interceptor^®^ G2 and Royal Guard^®^) in comparison to control nets (standard Interceptor^®^) collected from the cRCT community at different time points across their operational lifespan. Our objective was to understand how they compare with the entomological outcomes results of the trial.

## Methodology

### Experimental hut study site

The experimental hut (EH) study was conducted in the Magu district Mwanza, Tanzania, specifically in the Welamasonga village (2°34.673’ 33°07.170’) between 2020 to 2022. In this village, six experimental huts were constructed in the north part of the Misungwi cRCT area (16) (see Figure 1). A detailed description of the cRCT area can be found in the protocol (12).

**Figure 1:**
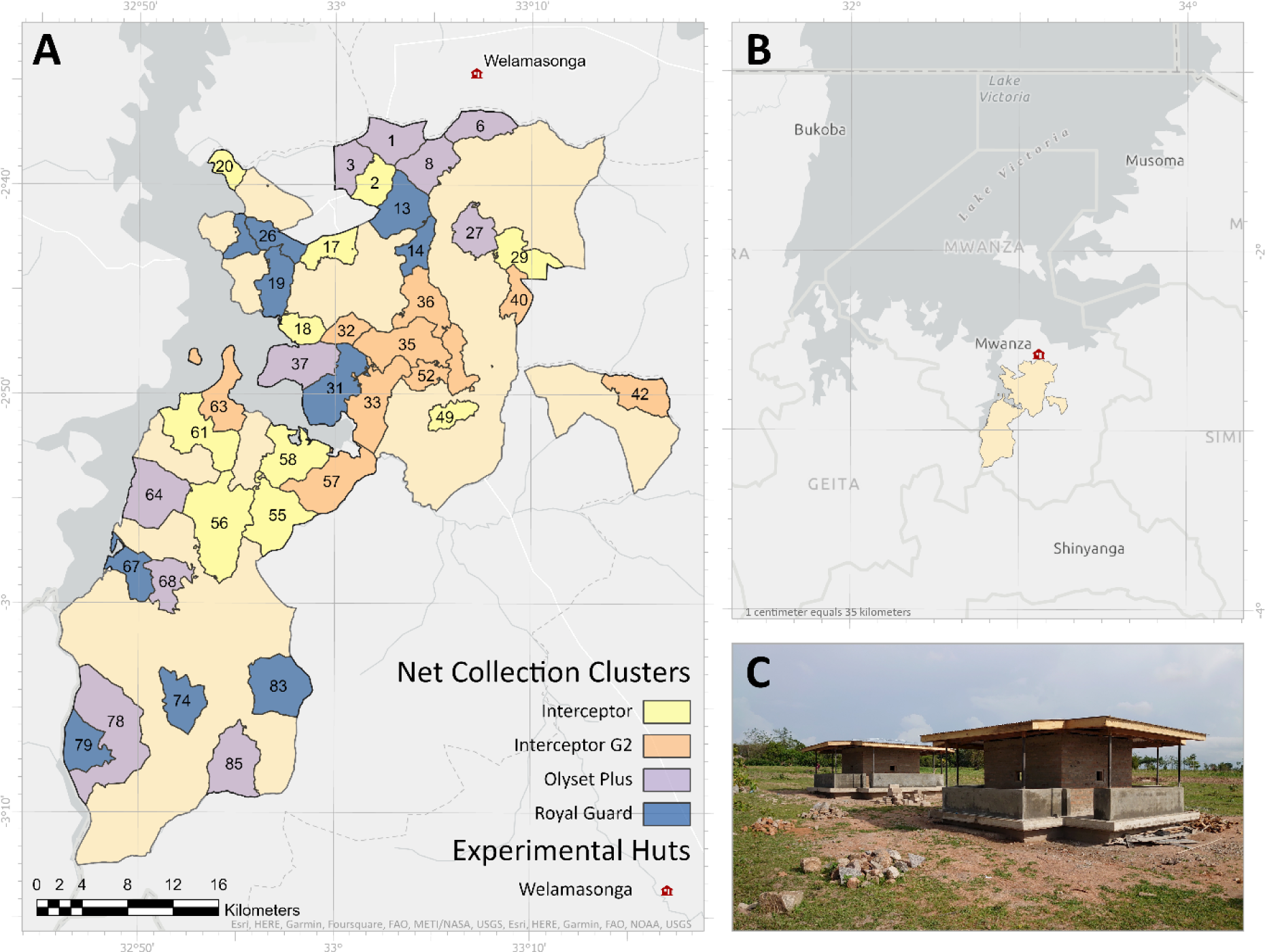
Map showing the location of experimental huts in the north part (B &C) of the main cRCT area and the clusters where the net was collected (A).

Magu experiences the same climatic conditions as the cRCT site, characterized by two rainy seasons (March – May and October – December) separated by a long dry season (June – August or September) and a short dry season (December or January – February) (16). The area surrounding the experimental huts consists of open fields used for cultivating rice and vegetables, with irrigation from ponds. The main vector species in this area are *An. funestus* sensu stricto (s.s), *An. arabiensis* and *An. gambiae* sensu stricto (s.s). The composition of these species varies with the season, with *An. gambiae* s.l dominating during the rainy season and *An. funestus* s.l. during the dry season.

Baseline resistance monitoring reported 69% of *An. gambiae* s.l. exposed to the diagnostic dose of alpha-cypermethrin and 12% exposed to permethrin exhibited 24-hour mortality. As for *An. funestus*, only one insecticide was tested (permethrin using CDC bottle assay), and it resulted in a 25% mortality rate 24 hours after exposure to the diagnostic dose.

### Net cohort enrolment and withdrawal for experimental hut trials

The nets to be rotated in the experimental hut were collected from Misungwi district, Mwanza, Tanzania where the cRCT was conducted. The cRCT spanned between 2019 and 2022 and aimed to assess the effectiveness of two dual-AI LLINs and a pyrethroid-PBO LLIN, compared to standard pyrethroid-only nets against malaria infection (12). The four LLINs under evaluation were 1/ Royal Guard,^®^, a net combining pyriproxyfen, which is known to disrupt female reproduction and fertility of eggs, and the pyrethroid alpha-cypermethrin;; 2/ Interceptor^®^ G2, a mixture net incorporating two adulticides with differing modes of action; chlorfenapyr (a pyrrole) and a pyrethroid (alpha-cypermethrin;); 3/ Olyset^TM^ Plus, an LLIN which incorporates a synergist PBO, to enhance the potency of pyrethroid insecticides and; 4/ Interceptor^®^, an alpha-cypermethrin-only LLIN and the positive control reference intervention (Table 1).

**Table 1:** EHT treatments evaluated.

| Treatment | Status of the net | Number of nets |
| --- | --- | --- |
| Olyset™ Plus | Field collected | 30 nets (tx*) |
|  | New unwashed with 6 holes | 6 nets (t0**) |
| Interceptor® G2 | Field collected | 30 nets (tx) |
|  | New unwashed with 6 holes | 6 nets (t0) |
| Royal Guard® | Field collected | 30 nets (tx) |
|  | New unwashed with 6 holes | 6 nets (t0) |
| Interceptor® | Field collected | 30 nets (tx) |
|  | New unwashed with 6 holes | 6 nets (t0) |
| Positive control | New Interceptor® with 6 holes | 6 nets (t0) |
| Negative control | New untreated with 6 holes | 6 nets (t0) |

\*tx are net collected at either 6, 12, 24, 30 or 36 months.
\*\* new unwashed net

In January 2019, study nets were distributed across the study arms in 84 clusters within the study area. One month after distribution, 1950 nets in 5 clusters per treatment arm were labelled with a unique number. Thirty of these labelled nets were collected at intervals 6, 12, 24, 30 and 36 months for use in EHT. The sampling design is explained in more detail elsewhere (16). Throughout the daily use of the nets, natural holes, wear and tear developed, and these were measured using a hole template. This measurement aimed to estimate the total hole area in each net brand, followingthe WHO guidelines (14). The nets were categorized based on the total area: “good” if the area was less than 79 cm^2^, “damaged” if the hole area ranged from 80 to 789cm^2^, and “torn” if the hole area was greater than 790 cm^2^ (17).

Before distribution, six new nets from each brand were retained to be tested in the experimental huts. To minimize the effect of net withdrawal, each collected net was replaced by a new net of the same type. However, these replacement nets were not included in the study. Households remained part of the eligible cohort until no enrolled nets were available.

### Experimental hut study

Nets collected at each time point were assessed in the same year, with those collected after 12 months of use were tested in 2020, those collected after 24 months in 2021, and those collected after 36 months in 2022. To account for vector seasonality and estimate net efficacy against different *Anopheles* species, four hut studies were conducted for nets collected at each time point over a year (16).

Each EHT study was done over a 6-week period using six experimental huts with the treatment presented in table 1. Sleepers were rotated between huts on successive nights to account for individual attractiveness, and treatments were rotated every week following a random Latin square design. For each hut trial, during the 36 day/nights, each net (30 tx and 6 t0) from each treatment (Olyset^TM^ Plus, Interceptor^®^ G2, Royal Guard^®^ and Interceptor^®^) was rotated every night. For all new intact LLINs, six holes of 4 x 4 cm were cut following WHO guidelines.

In each hut, cloth sheets were laid on the floor and sugar solution was provided at night in the window traps to reduce mosquito mortality (18). The day after net installation in the hut, mosquitoes were collected using standard mouth aspirators from the floor, inside nets and on walls and ceilings. The mosquitoes were packed in paper cups labelled with the collection date, hut number, net type, collection area (exit/room/net) and collector initials. All mosquitoes were sent to the National Institute for Medical Research (NIMR) insectary. Mosquitoes were identified morphologically to species (19) and categorized by gonotrophic status (i.e., unfed, freshly fed, semi-gravid and gravid).

Mosquito mortality was monitored every day up to 72 hours in a controlled environment. The effect of pyriproxyfen on reproductive outcomes was assessed on mosquitoes collected from the huts deployed with Royal Guard^®^, Interceptor^®^ and untreated nets by dissecting gravid *Anopheles* 72 hours after collection when eggs should normally have fully matured (20, 21).

After a 72-hour holding period, *An. gambiae* s.l. were further identified to species-level using a TaqMan PCR assay to distinguish *An. gambiae* s.s. from *An. arabiensis* (22) and the same was done for *An. funestus* s.l. to distinguish between *An. funestus* s.s and *An*. *parensis* (23). Half of the blood fed and unfed, alive and dead mosquitoes were packed in RNAlater^®^ for determination of species and cytochrome P450 expression levels using qPCR (24, 25).

### Resistance monitoring

Wild *An. gambiae* s.l and *An. funestus* s.l were collected from houses adjacent to the experimental hut site around 6:00 am to 7:00 am in parallel of EHT study. Mosquitoes were morphologically identified to species and kept for three days to allow digestion of blood meal before bioassay testing.

Resistance intensity to the insecticides contained in each LLIN was assessed using WHO/CDC bottle bioassays every year. Mosquitoes were exposed to the diagnostic doses of alpha-cypermethrin or permethrin and increased to 2, 5 and 10 times of diagnostic concentration for 30 minutes, or chlorfenapyr (100 μg/ml), pyriproxyfen (100 μg/ml) for 60 minutes; PBO pre-exposure was also performed using WHO tube bioassays followed by CDC exposure to pyrethroid.

### Outcome measures

Primary outcomes were 1/ 72-hour mortality for Interceptor^®^ G2, 2/ 24-hour mortality for Olyset^TM^ Plus and 3/ fertility for Royal Guard^®^, calculated as the proportion of blood fed females alive at 72 hours with fully mature eggs. Secondary outcomes were 1/ 24-hour mortality for Royal Guard^®^ and for all the nets 2/ blood feeding: proportion of blood fed mosquitoes collected, 3/ deterrence: percentage reduction in density of *Anopheles* in treatment huts compared to negative control huts (fitted with untreated nets), 4/ exophily: proportion of mosquitoes that exited early and were found in exit traps compared with the untreated control huts.

### Data analysis

Data analysis was performed using STATA software version 17. After data cleaning, four nights of collections were removed due to reporting errors. Descriptive statistics were used to estimate the proportion of mosquito species collected each year.

Mixed effect generalized linear models with logit link function were used to compare proportional outcomes (mortality, blood feeding and fecundity) of Interceptor^®^ G2, Royal Guard^®^ and Olyset^TM^ Plus to reference Interceptor^®^ net (pyrethroid only field collected). Each models included treatment, hole index and net age as fixed effect and hut, sleeper and week as random effect to account for variation between sleepers, huts, collection weeks and seasonality. The interaction between treatment and net age was reported using odds ratios with their 95% confidence intervals (CI) and p-values were used to assess statistical significance at the 0.05 level for comparisons of mortality and blood-feeding. Mortality and blood feeding graphs were plotted using ggplot in R software.

For resistance data, lethal dose values (LD25, LD50, LD95 and LD99) were calculated using a probit model with log-10 transformed data in IBM SPSS v28 software. The curve estimation was based on the probability of mosquito death as a function of the total number of mosquitoes and insecticide dose (26). Point estimates of LDs and 95% CIs were then back-transformed to their original scale to obtain the reported values, indicating the difference in diagnostic dose of an insecticide required to kill 25%, 50%, 95% or 99% of tested mosquitoes. Comparisons of LD50 values among clusters and/or years were statistically estimated using the Relative Median Potency (RMP), calculated as the ratio of point estimates with simultaneous 95% CIs. Comparisons of “potency” in this context are median lethal concentrations/doses. A ratio of “1” is considered insignificant, meaning LD50 was equal among comparison groups. Reduction in fertility was calculated as ((proportion fertile in control-proportion fertile in treatment)/ proportion fertile in untreated net control)*100.

## Results

### Species composition and outcomes in negative and positive control huts over the 3 years

A total of 12 experimental hut trials were conducted between 2020 and 2022, resulting in 2,588 collection nights. During this period, 17,040 male and female mosquitoes were collected with 87% (14,841/17,040) of them being female. Among the female mosquitos, 26% (3,925/14,841) were identified as *Anopheles,* while the remaining were *Culex quinquefasciatus*.

For all collection years combined, 63.4% (2488/3925) of the *Anopheles* were identified as *An. gambiae* s.l. while the remaining 36.6% (1437/3925) were *An. funestus* s.l.. Among the *An. gambiae* complex identified to species-level, 56.8% (673/1185) were determined to be *An. gambiae* s.s, while the rest were classified as *An. arabiensis*. Notably, the proportion of *An. gambiae* s.s. was the highest in 2020 and subsequently decreased in years (as shown in Table 2).

**Table 2:** Hut trials entomological characteristics (density and species composition) over the three years of follow-up.

|  | Year 1: 2020 |  | Year 2: 2021 |  | Year 3: 2022 |  |
| --- | --- | --- | --- | --- | --- | --- |
| Total hut/night collection | 863 |  | 864 |  | 864 |  |
| Total mosquitoes collected (female) | 4320 (3869) |  | 7424 (6612) |  | 5296 (4360) |  |
| Total *Anopheles* collected (female) | 1974 (1611) |  | 1904 (1457) |  | 1204 (857) |  |
| *Anopheles* vectors: N, mean [95% CI] | 1611 | 1.9 [1.7 - 2.0] | 1457 | 1.7 [1.5 - 1.8] | 857 | 0.9 [0.9 - 1.1] |
| *Culex* species: N, mean [95% CI] | 2270 | 2.6 [2.4 - 2.8] | 5155 | 5.9 [5.2 - 6.7] | 3503 | 4.1 [3.7 - 4.4] |
| **Species composition n/N, % [95% CI]** |  |  |  |  |  |  |
| *An. gambiae* s.l./*Anopheles* | 1161/1611, 72.1%, [68.9 - 75.1] |  | 842/1457, 57.8% [51.9 - 63.4] |  | 485/857, 56.6% [50.4 - 63.5] |  |
| *An. funestus* s.l./*Anopheles* | 450/1611, 27.9% [24.4 - 30.7] |  | 615/1457, 42.2% [36.6 - 48.1] |  | 372/857, 43.4% [36 - 49.6] |  |
| *An. gambiae* s.s/*An.gambiae* s.l | 347/410, 85.0% [80.7 - 87.8] |  | 181/334, 54.0% [ 48.8 - 59.4] |  | 143/435, 33.0% [29 - 37] |  |
| *An. funestus* s.s/*An.funestus* s.l | 151/153, 99.0% [ 94.8 - 99.6] |  | 436/460, 95.0% [ 92.3 - 96.5] |  | 321/368, 87.0% [83 - 90] |  |

*Anopheles* mortality for untreated nets (negative control huts) was less than 5% after 24 hours which meets the WHO recommended threshold, while the exophilic rate ranged between 62% and 68% depending on the year. Blood feeding was between 18% and 21% in huts fitted with a new standard pyrethroid LLIN Interceptor^®^ (positive control huts); 24-hour mortality was low and ranged from 8% to 12%. The mortality against *An. funestus* complex for the whole period of study varied between 4% to 10% while it ranged between 7% to 18% for *An. gambiae* (see additional file 1).

### EHT results on mortality and blood feeding

There was higher 24-hour mortality in *Anopheles* collected in huts fitted with new (0 month) Olyset^TM^ Plus compared to a new standard pyrethroid LLIN (raw data 38% vs 6%), adjusted OR: 13.6, 95% CI: 4.4 – 41.3, p-value < 0.001 (Figure 2). Mortality in Olyset^TM^ Plus dropped over time and while it remained slightly higher compared to standard pyrethroid LLINs of the same age, the difference was not significant after nets were used for 12 months or more (12 months: 17% vs 13%; OR: 2.1, 95% CI: 1.0 – 4.3; p = 0.112, 24 months: 12% vs 11%, OR: 1.4, 95% CI: 0.6 – 3.3; p = 0.310 and 36 months: 10% vs 7%, OR: 1.0, 95% CI: 0.3 – 3.5; p = 0.890) (see Figure 2 & additional file 2a). By species, 24-hour mortality remained significantly higher in Olyset^TM^ Plus compared to the standard pyrethroid LLIN in *An. gambiae* s.l (23% vs 13%, OR: 2.6, 95% CI: 1.0 – 6.4; p = 0.045) at 12 months but no effect was observed on the *An. funestus* complex at this time point (see additional file 2b). Blood feeding for both species was lower in Olyset^TM^ Plus compared to Interceptor^®^ LLINs up to 12 months but the difference was not significant at any of the following time points (see Figure 4).

**Figure 2:**
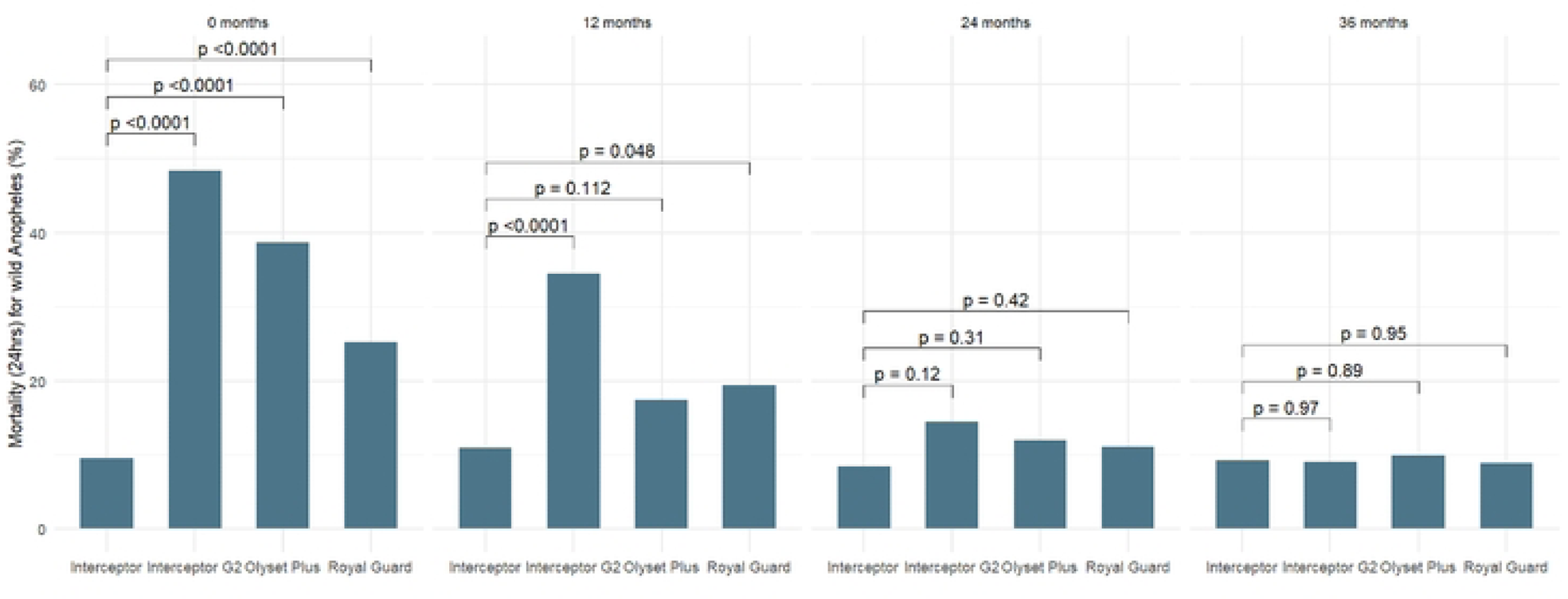
Model output mortality (24hrs) of wild free flying female *Anopheles* in experimental hut by net type and age.

Interceptor^®^ G2 LLINs provided higher 72-hour mortality compared to Interceptor^®^ LLINs when new (raw data 58% vs 14%), (OR: 11.9, 95% CI: 4.8 – 29.7 p < 0.001) and after 12 months of use (43% vs 21%, OR: 3.5, 95% CI: 1.9 – 6.6, p < 0.001) (see Figure 3 & additional file 2a). The effect was observed for both *An. gambiae* s.l. and *An. funestus* (see additional file 2b and 2c). At 24 and 36 months, the difference in mortality was no longer significantly higher. Similar findings were observed for 24-hour mortality (see Figure 2). In terms of blood feeding inhibition, there were no differences between Interceptor^®^ G2 compared to Interceptor^®^ LLINs at any time point (see Figure 4).

**Figure 3:**
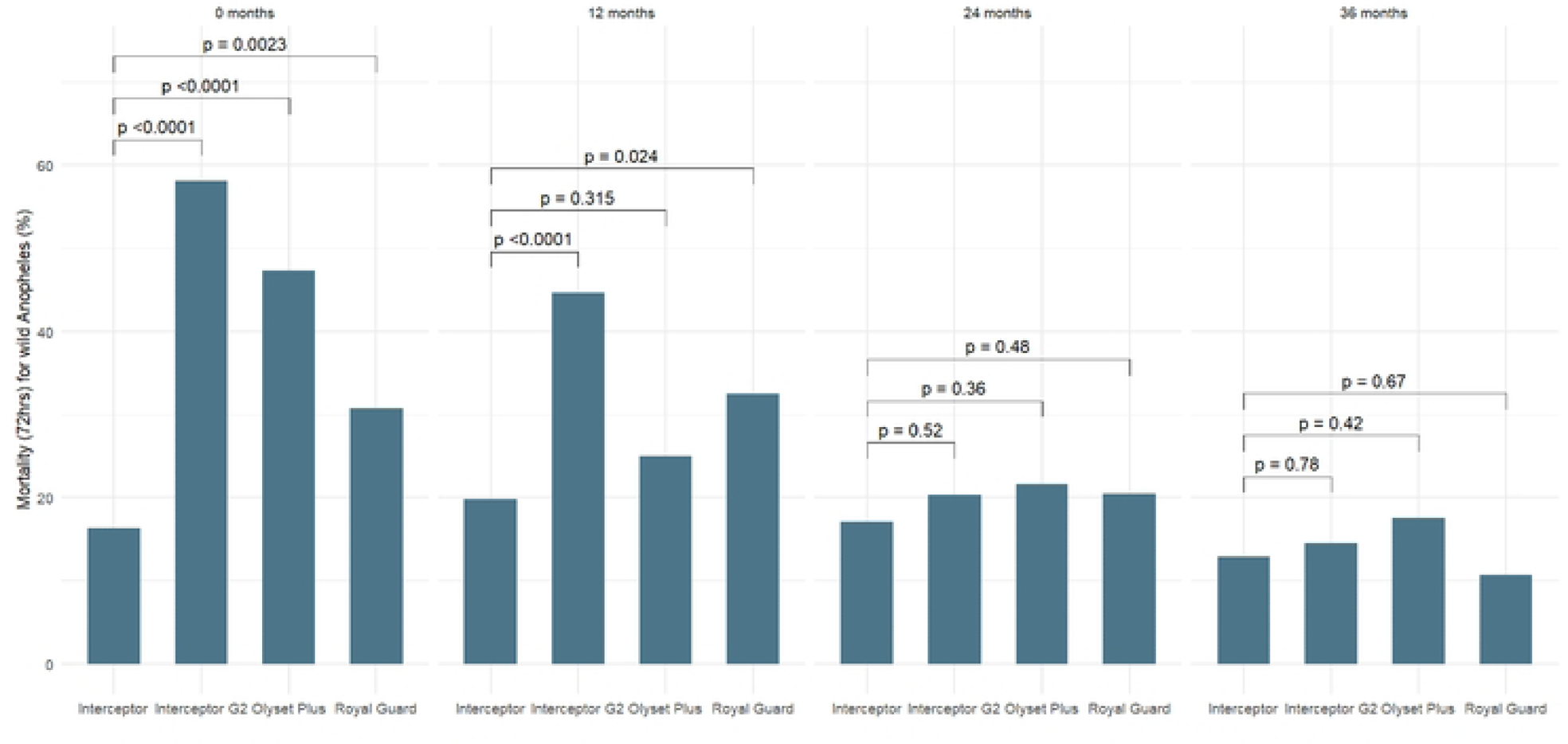
Model output mortality (72hrs) of wild free flying female *Anopheles* in experimental hut by net type and age.

**Figure 4:**
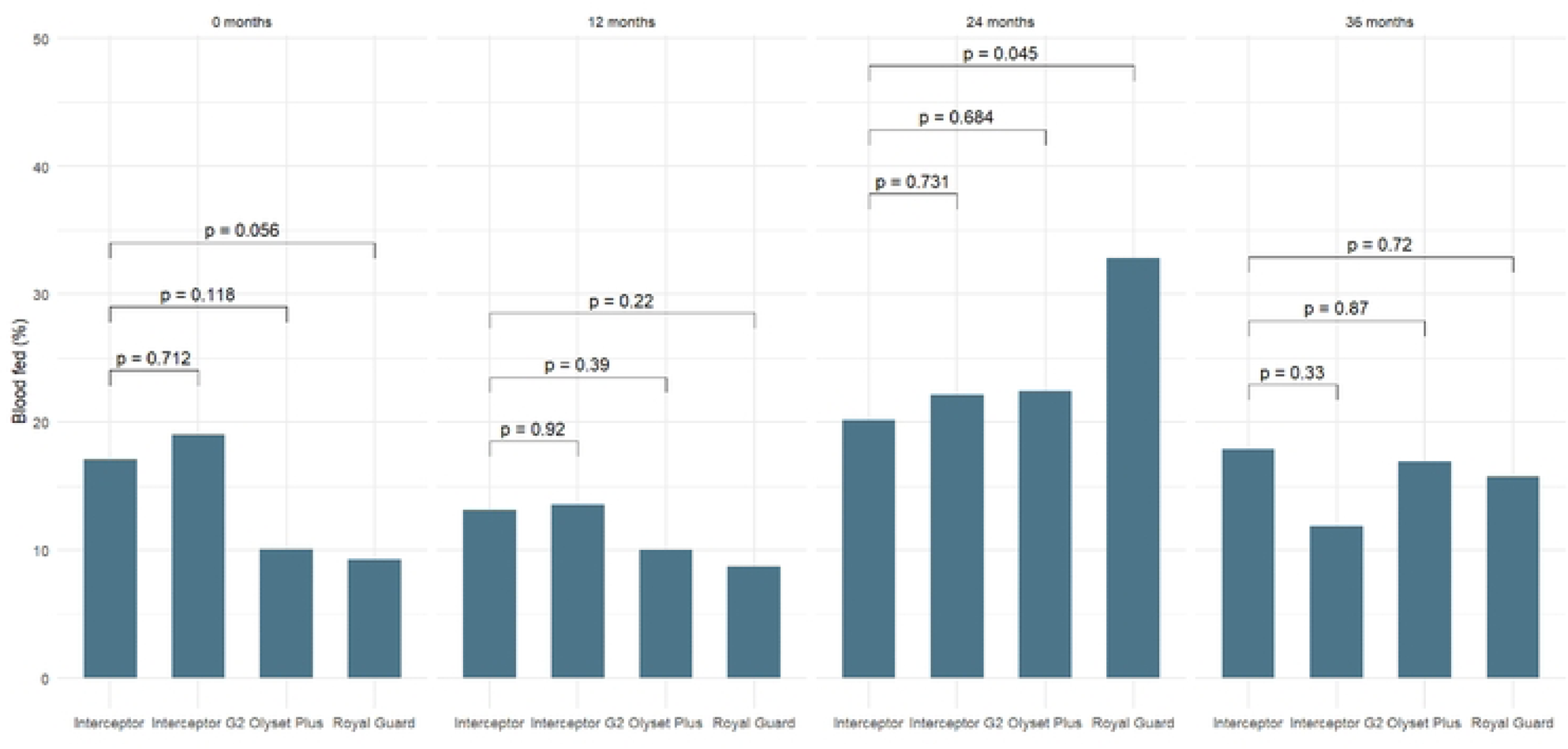
Model output blood feeding in wild free flying female *Anopheles* in experimental hut by net type and age.

For Royal Guard^®^, the primary outcome was effect on fertility. Fertility rate varied between 94% and 100% for *Anopheles* collected from untreated and standard pyrethroid Interceptor^®^ LLINs. In Royal Guard^®^ LLINs huts, a total of 707 female *Anopheles* were collected, of which 138 were blood fed and only 91 alive after 72 hours and available for dissection (see Table 3). Fertility was slightly reduced when Royal Guard^®^ was new (88%) or after 12 months of use (75%). At 24 and 36 months, all mosquitoes were found with fully mature eggs and categorized as fertile. Interestingly, despite small numbers being dissected, a reduction in fertility was only observed for *An. gambiae* s.l and not *An. funestus*. Mortality outcomes were also monitored and were found significant at 0 months and 12 months compared to Interceptor^®^ LLINs (additional file 2a). Blood feeding was lower at 0, 12 and 36 months in Royal Guard^®^ compared to standard pyrethroid LLINs but the difference was not significant and decreased with time, while at 24 months more blood fed *Anopheles* were found in the Royal Guard^®^ huts (as shown in Figure 4 and Table 3) than in standard pyrethroid LLINs.

**Table 3:**
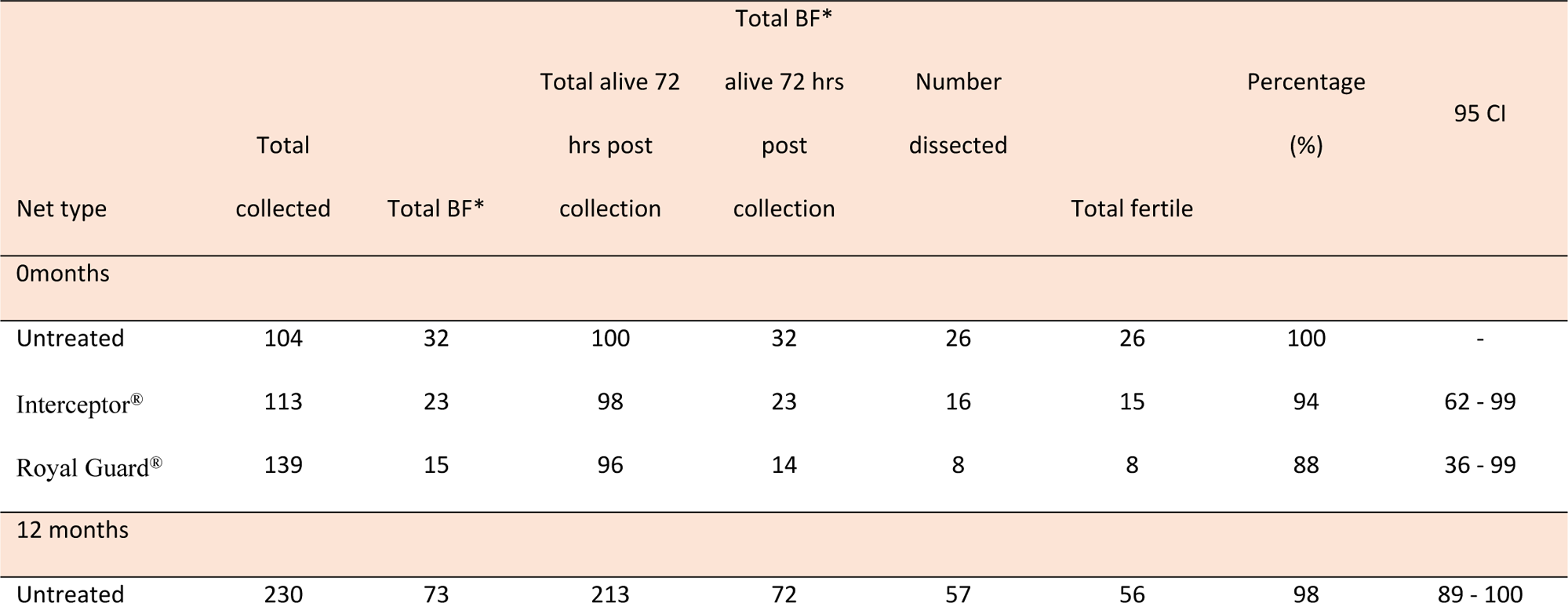
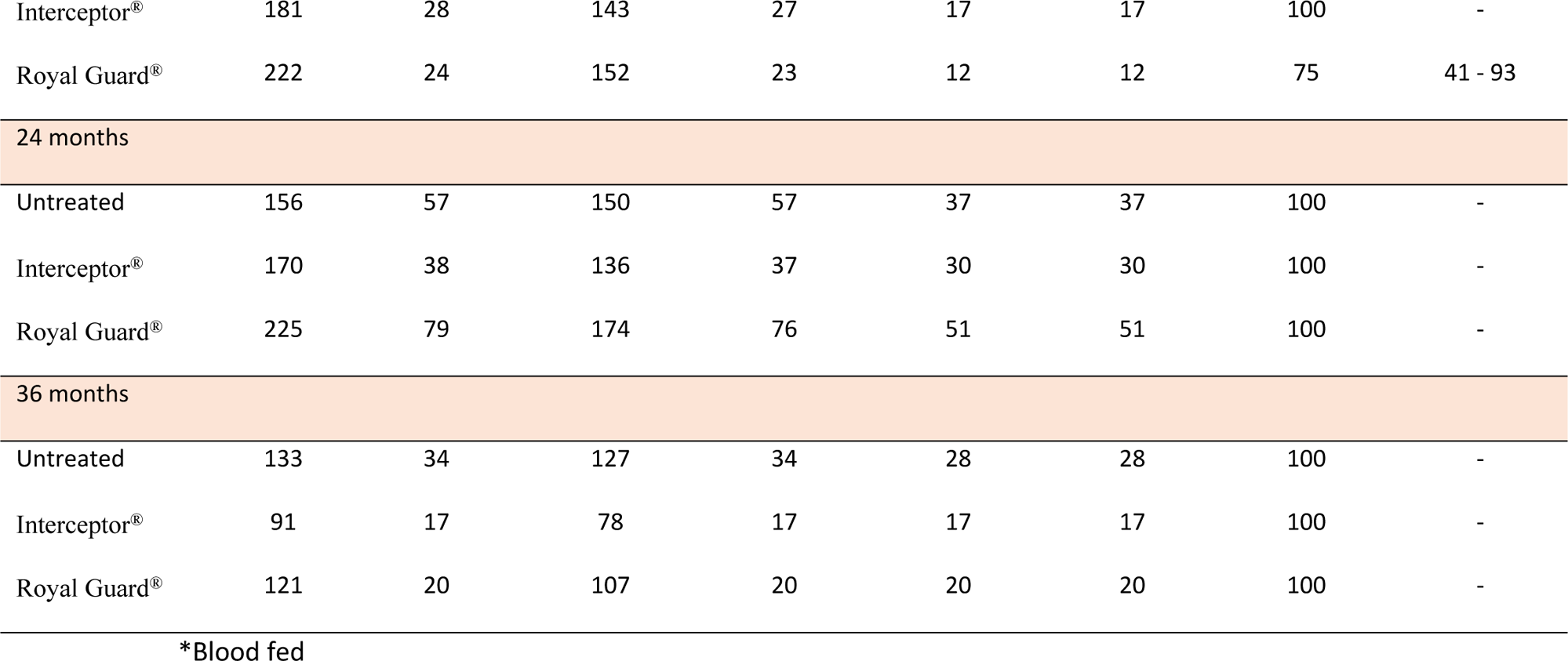
Fertility in wild free flying female *Anopheles* in experimental hut fitted with Interceptor^®^, untreated net and Royal Guard^®^ by age.

### EHT results on deterrence, exit and net penetration

There was no clear pattern with deterrence for *An. gambiae* s.s.. For nets aged 0 and 24 months there was no deterrence effect in the treatment huts relative to huts fitted with untreated nets while at 12 and 36 months there was some reduction of *An. gambiae* s.s. entered in huts with treated nets (additional file 3). For *An. funestus*, regardless of net age there was some hut entry reduction in huts fitted with Interceptor^®^ G2 and standard pyrethroid Interceptor^®^ LLINs while that was not observed at any time point for Olyset^TM^ Plus and Royal Guard^®^ (except at 36 months for the latter). Overall, the highest exit rate for *Anopheles* was found in treatment huts compared to untreated huts, but there were no significant differences between Interceptor^®^ G2, or Royal Guard^®^, or Olyset^TM^ Plus compared to Interceptor^®^ of the same age. Overall, penetration of nets by *Anopheles* was consistently lower for all the insecticide treated nets compared to untreated nets (additional file 3).

### Hole characteristics of nets sampled from the community and effect of hole and net age on vector blood feeding

The mean hole area for Interceptor^®^ G2 and Olyset^TM^ Plus LLINs aged 12 months for the good category was 26.5 cm^2^ and 27.9 cm^2^, respectively while the mean hole area in Royal Guard^®^ was much lower (16.2 cm^2^). On average, hole size increased with net age (see additional file 4). Overall, mortality and blood feeding observed was not impacted by the size or number of holes in any of the nets.

### Insecticide resistance in malaria vectors

Longitudinal changes in vector insecticide resistance intensity were assessed to determine their influence on vector mortality in the EHTs. Permethrin resistance intensity was high in *An. funestus* in year one LD50 = 292.9 [52.7 – 3906.3] but diminished over time (year two LD50 = 33.7 [20.5 – 81.7] and year three LD50 = 19.1 [13.5 – 36.2]). By comparison, levels of alpha-cypermethrin resistance intensity in *An. funestus* increased significantly during the EHTs (year two LD50 = 22.5 [12.9 – 65.1] and year three LD50 = 181.6 [61.8 – 1499.9]). In *An. gambiae* s.l., a similar decline in permethrin resistance intensity was evident (year one LD50 = 3417 [4.0 – 7319224.4], year two LD50 = 24.1 [15.5 – 48.3] and year three LD50 = 43.6 [26.4 – 100.0]). However, there was no parallel increase in alpha-cypermethrin resistance (year one LD50 = 0.24 [0.0 – 0.6], year two LD50 = 0.52 [0.2 – 0.9] and year three LD50 = 0.79 [0.3 – 1.3]). In both vector species complex, there was limited change in permethrin resistance intensity following PBO pre-exposure.

Following chlorfenapyr exposure, high 72-hour mortality was evident in both species complexes at year one (92% [95% CI: 87 – 97] and 91% [95% CI: 87 – 96] for *An. gambiae* s.l and *An. funestus* s.l., respectively); complete (100%) mortality was observed for both *An. gambiae* s.l and *An. funestus* s.l, in year two and three.

During pyriproxyfen testing, there was no impact on fertility in *An. funestus* s.l in year one while an 8.1% [8/98] reduction was observed in *An. gambiae* s.l. in year one. For year two and three, only *An. gambiae* s.l were tested and 8.8% [5/57] and 6.7% [1/15] reduction in fertility was observed respectively. Overall, sterility effect was less than 10%.

Dynamic changes in expression of eight metabolic genes (CYP6P4, CYP6Z1, CYP4G16, CYP9K1, CYP6M1, CYP6P1, CYP6P3 and GSTF2) were observed in PCR-confirmed *An. gambiae* s.s. between trial years one and two (Figure 5). CYP9K1, CYP6M2 and GSTE2 displayed consistent minimal over-expression relative to the susceptible colony strain. By comparison to the untreated net arm, significant declines in CYP6P4 expression were observed in Interceptor^®^ (washed and unwashed) and Olyset^TM^ Plus exposed mosquitoes and no significant decrease in CYP6Z1 expression was present in Royal Guard^®^ and Interceptor^®^ G2 exposed mosquitoes between years one and two; while a significant increase in CYP6P1 expression was apparent in vectors collected from Royal Guard^®^ EHs.

**Figure 5:** Gene expression in wild field collected *An.gambiae* s.l relative to colony susceptible population over two years in untreated net (A), Interceptor^®^ G2 (B), standard Interceptor^®^ (field collected) (C), standard Interceptor^®^ (washed once) (D), Royal Guard^®^ (E) and Olyset ^TM^ Plus (F). Error bars represent 95% confidence intervals. Statistically significant differences in expression levels between trial years are indicated as follows: ns=not significant; *= p-value < 0.05; ** = p-value < 0.01; ***= p-value < 0.001

## Discussion

These series of experimental hut trials were conducted to assess the impact of interventions (dual-AI LLINs) on mortality and blood feeding rates in resistant *An. gambiae* s.l and *An. funestus* s.l. vector populations, and to better understand the associated cRCT outcomes. When new, Interceptor^®^ G2 and Olyset^TM^ Plus provided significantly higher killing effect compared to the reference net (Interceptor^®^) against wild resistant *Anopheles* found in Magu district. Mortality was also generally higher for these two nets at 12 months, but there was only a strong evidence for a difference for Interceptor ^®^ G2. Royal Guard^®^ did not have a significant impact on fertility outcomes at any of the time points.

The highest effect of Interceptor^®^ G2 on vector mortality was observed for both vectors *An. gambiae* s.l. and *An. funestus* up to 12 months, with no observable difference after 24 and 36 months of net use compared to Interceptor. Previous experimental hut trials done in Tanzania (27) assessing new unwashed Interceptor^®^ G2 reported a 72-hour mortality around 50% against *An.funestus* similar to our findings, while mortality was in general higher in Benin (71%) and in Cote d’Ivoire (87%) against *An. gambiae* s.l.. Interestingly, in these EHTs, after 20 washes (supposed to mimic a 36 month field net) 72-hour mortality was 52% in Tanzania, 65% in Benin (11) and 82% in Cote d’Ivoire (28), all much higher than what we found in our EHT at any given time point. In Cameroon, Interceptor^®^ G2 has been reported to induce higher mortality than a standard net after 20 washes against the *An. funestus* complex (29). In our study, the insecticidal content over time depleted quicker compared to those exposed to laboratory washing procedures; indeed, the residual concentration of chlorfenapyr was only 8% of the initial content after 36 months of operational use while the chlorfenapyr retention observed in nets washed 20 times were 32% in Tanzania (27) and 37% in Benin (11, 27)

Overall mortality induced by field collected Olyset^TM^ Plus LLINs was significantly higher than pyrethroid-only net (Interceptor^®^) when the nets were new but at no other time point. When looking at the effect per species mortality was also significantly higher in *An.gambiae* s.l. at 12 months which was not the case for *An.funestus*. In the cRCT conducted as part of this study (12), Olyset^TM^ Plus LLIN was only effective for a year while a previous cRCT conducted in Muleba-Tanzania showed that Olyset^TM^ Plus LLIN arm had lower malaria than Olyset^TM^ Net (8) and provided personal protection (30) up to 21 months of use in an operational setting. The EHT results from the present study suggest that Olyset^TM^ Plus might control *An.gambiae* s.s. better than *An.funestus*, which could explain the difference between those two cRCTs as in Misungwi the main vector was *An. funestus* (31) while in Muleba it was *An. gambiae* s.s. (8). The efficacy of Olyset^TM^ Plus decreased with net age and this correlates with the depletion of permethrin and PBO content after 36 months of community use (8.3g/kg vs 20g/kg when new and ∼0.7g/kg vs 10g/kg respectively). Similar findings were reported in Uganda (32) with low retention (3.7g/kg) of PBO content, 25 months post distribution for Olyset^TM^ Plus LLIN and Kenya (33) 87% of PBO and 52% of permethrin were lost after 36 months of community use, while PBO retention was higher when nets were washed 20 times in the laboratory (2.0g/kg) in Benin (34).

In Royal Guard^®^, no significant sterility effect was observed through the 3 years with only 33.3% of the *Anopheles* considered sterile after exposure to new unwashed Royal Guard^®^ nets and 25% after 1 year of use. No sterility effect was observed at 24 and 36 months. Interestingly, the impact of Royal Guard^®^ on mortality was observed up to 12 months, in addition the limited number of blood fed *Anopheles* still alive at 72 hours, suggest that the combination of pyrethroid and pyriproxyfen may have had more impact on mortality and blood feeding than fertility for the first year. A study in Benin (10) using Royal Guard^®^ LLINs showed reduction in the reproduction ability of mosquitoes up to 20 times washes in a laboratory setting however the effect was lower (25%) in experimental huts. Laboratory evaluation of another brand of pyriproxyfen net, Olyset Duo, conducted in Moshi-Tanzania against *An.gambiae* s.l. (35) reported 34.6% fertility after being exposed in tunnel tests after 20 washes (35). However, entomological assessment of this brand in a cRCT in Burkina Faso showed that the sterility effect was only observed for one month after LLIN distribution (36). The difference in performance between laboratory and community studies could be explained by more stringent washing methods and abrasion during daily use. In our study only 28% of pyriproxyfen and 62% of alpha-cypermethrin remained on the nets after 36 months while insecticide retention was higher when washed in the laboratory (57% and 76% for pyriproxyfen and alpha-cypermethrin, respectively)(10). To prequalified new LLIN products, WHO recommends different phases of evaluation including phase II laboratory wash resistance which is assumed to corelate with 36 months of community use (14). However, this study reveals that wash resistance (20 times) does not correlate with the 36 months LLIN. The WHO could review guidelines for evaluation (phases I, II and III) and increase number of washes and recommend abrasive washing method to mimic 36 months LLIN in operation setting.

Resistance intensity monitoring demonstrated high resistance to permethrin in *An. funestus* in the first year, which was lost in the second and third year. By comparison, alpha-cypermethrin resistance intensity increased significantly over trial years in *An. funestus,* which aligns with resistance monitoring results from the main cRCT (26). This may be due to an increase in the proportion of zoophilic *An. arabiensis* recorded resting in the huts, (31).

Chlorfenapyr demonstrated high killing effect in all three years of resistance monitoring without evident selection whilepyriproxyfen, failed to induce significant sterility effect against *An. gambiae* s.l and *An. funestus* complex throughout the same period. Pyriproxyfen resistance results is consistent with those observed in cRCT study site and could explain the relatively small effect observed in both cRCT and EHT presented (26). The molecular analysis for the detoxification enzymes revealed overexpression of genes (CYP6P3) in Interceptor^®^, Interceptor^®^ G2 and Olyset^TM^ Plus which strongly metabolize pyrethroid insecticide, as it was observed in the study examining the functional genetic keys that confer resistance in African malaria vectors, *An.gambiae* s.l. (37–39).

One of the limitations in this study was the variation in species composition over time which could explain why some of the nets did not perform as anticipated; future studies may evaluate nets of different ages side-by-side to control for this heterogeneity across years. A second limitation is that a relatively small number of blood fed *Anopheles* alive at 72 hours were available for dissection and therefore the impact of Royal Guard^®^ on sterility might have been slightly underestimated.

Overall, the reduction in efficacy of Olyset^TM^ Plus and Royal Guard^®^ LLINs observed in the EHTs seems to match the entomological and epidemiological findings in the cRCT conducted in Misungwi with Olyset^TM^ Plus providing superior protection against malaria infection and vectors outcomes compared to Interceptor up to 12 months only (12) while there was no significant differences for Royal Guard^®^. However this was less clear for Interceptor^®^ G2. While in the current EHTs Interceptor^®^ G2 outperformed Interceptor^®^ only up to 12 months, the cRCTs outcomes reported significant malaria prevalence reduction and vector densities in the Interceptor^®^ G2 arm compared to standard LLIN at 24 months (12, 13) and even up to 36 months (15). Differences in species composition in the EHT and cRCTs area could explain the difference. Indeed, there was a majority of *An.funestus* found in the cRCTs while *An.arabiensis* was the predominant species in the EHT when the 24 and 36 months nets were tested. Matowo et al. (31) have reported that Interceptor^®^ G2 was less effective against exophilic (*An. arabiensis)* mosquitoes. Chlorfenapyr is a complex insecticide, and a recent study reported potential additional properties to theHowever, we also found that Interceptor^®^ G2 superior killing effect that impacts directly on malaria against *An.funestus* was observed up to 12 months. Differences between cRCT and EHT outcomes may be explained by potential effect of chlorfenapyr on parasite development in mosquitoes (40), which could explain the stronger and longer effect observed in the community. What the result of the present EHT also highlighted is that the reduction in effect observed in the cRCT in Tanzania over the three years might not only be due to reduction in net usage but also to the sharp decrease in partner AI. or PBO resulting in lower killing effect as the net aged.

Modelling of EHT data have been used to parameterize mechanistic models for malaria vectors and predict the epidemiological efficacy of LLINs (41). The use of EHT data which better correlates with cRCT outcomes as observed in the present study, especially for the quick acting and neuro acting LLINs, may improve the fit of these models and could be sufficient for the evaluation of second in class products.

## Conclusion

Olyset^TM^ Plus Royal Guard^®^ LLINs did showed superiority compared to Interceptor^®^ LLIN only when new and supported the findings of the cRCT reporting the limited effect in time against epidemiological and entomological outcomes. Interceptor^®^ G2 LLINs were superior against *An. gambiae* s.l and *An. funestus* complex, compared to Interceptor^®^ LLINs only up to 12 months while they provided superior protection against malaria for 3 years in an associated cRCTs. Further research is required to explore additional effects of chlorfenapyr in order to comprehend its full impact on malaria transmission.

## Data Availability

The data will be available upon publication. All data for this study are store in data repository and the access will be given for the particular dataset. Currently, there is still additional data analysis ongoing, and we are unable to share the data at the time of the review but data will be made available at the end of the project in the LSHTM repository.

## Acknowledgements

Special thanks to the community for allowing us to construct our experimental hut in their area, to the project technicians for collection, processing, and identification of mosquitoes, and to the volunteers for agreeing to participate in the study. We would like to thank Jackie Cook and David Macleod for preparing the random latin square stata do file, Chad Cross for the SPSS code for resistance analysis.

## Authors contribution

JLM contributed to supervise field work and drafted the manuscript; LAM contributed to analysis and revised the manuscript; NP was the trial principal investigator, participated in study design, implementation of field activities and revised analysis and manuscript; FM contributed to study design and supported the field activities;; JFW and AM contributed to study implementation; MR designed the study, implementation of field activities and contributed to writing and revising the manuscript; EB, SL, MK and PH contributed to data collection, sorting and storage of mosquitoes; BS, CS and OM performed molecular assays; CT and MK contributed to the analysis and map; NM supervised field work. All authors read and approved the final manuscript.

## Reference

1. WHO. Malaria report. World Health Organisation. 2021.

2. Ngufor C, N’Guessan R, Fagbohoun J, Todjinou D, Odjo A, Malone D, et al. Efficacy of the Olyset Duo net against insecticide-resistant mosquito vectors of malaria. Sci Transl Med. 2016;8(356):356ra121.

3. Tiono AB, Pinder M, N’Fale S, Faragher B, Smith T, Silkey M, et al. The AvecNet Trial to assess whether addition of pyriproxyfen, an insect juvenile hormone mimic, to long-lasting insecticidal mosquito nets provides additional protection against clinical malaria over current best practice in an area with pyrethroid-resistant vectors in rural Burkina Faso: study protocol for a randomised controlled trial. Trials. 2015;16:113.

4. Churcher TS, Lissenden N, Griffin JT, Worrall E, Ranson H. The impact of pyrethroid resistance on the efficacy and effectiveness of bednets for malaria control in Africa. Elife. 2016;5.

5. World Health O, Who Pesticide Evaluation Scheme. Working Group. Meeting 15th GS. Report of the fifteenth WHOPES working group meeting: WHO/HQ, Geneva, 18-22 June 2012: review of Olyset plus, Interceptor LN, Malathion 440 EW, Vectobac GR. Geneva: World Health Organization; 2012 2012.

6. Menze BD, Mugenzi LMJ, Tchouakui M, Wondji MJ, Tchoupo M, Wondji CS. Experimental Hut Trials Reveal That CYP6P9a/b P450 Alleles Are Reducing the Efficacy of Pyrethroid-Only Olyset Net against the Malaria Vector Anopheles funestus but PBO-Based Olyset Plus Net Remains Effective. Pathogens. 2022;11(6).

7. Toe KH, Müller P, Badolo A, Traore A, Sagnon N, Dabiré RK, et al. Do bednets including piperonyl butoxide offer additional protection against populations of Anopheles gambiae s.l. that are highly resistant to pyrethroids? An experimental hut evaluation in Burkina Fasov. Med Vet Entomol. 2018;32(4):407–16.

8. Protopopoff N, Mosha JF, Lukole E, Charlwood JD, Wright A, Mwalimu CD, et al. Effectiveness of a long-lasting piperonyl butoxide-treated insecticidal net and indoor residual spray interventions, separately and together, against malaria transmitted by pyrethroid-resistant mosquitoes: a cluster, randomised controlled, two-by-two factorial design trial. The Lancet. 2018;391(10130):1577–88.

9. WHO. Report of the 20th WHOPES Working Group meeting. Geneva WHO; 2017. Contract No.: WHO/HTM/NTD/WHOPES/2017.04.

10. Ngufor C, Agbevo A, Fagbohoun J, Fongnikin A, Rowland M. Efficacy of Royal Guard, a new alpha-cypermethrin and pyriproxyfen treated mosquito net, against pyrethroid-resistant malaria vectors. Sci Rep. 2020;10(1):12227.

11. N’Guessan R, Odjo A, Ngufor C, Malone D, Rowland M. A Chlorfenapyr Mixture Net Interceptor(R) G2 Shows High Efficacy and Wash Durability against Resistant Mosquitoes in West Africa. PLoS One. 2016;11(11):e0165925.

12. Mosha JF, Kulkarni MA, Lukole E, Matowo NS, Pitt C, Messenger LA, et al. Effectiveness and cost-effectiveness against malaria of three types of dual-active-ingredient long-lasting insecticidal nets (LLINs) compared with pyrethroid-only LLINs in Tanzania: a four-arm, cluster-randomised trial. Lancet. 2022;399(10331):1227–41.

13. Accrombessi M, Cook J, Dangbenon E, Yovogan B, Akpovi H, Sovi A, et al. Efficacy of pyriproxyfen-pyrethroid long-lasting insecticidal nets (LLINs) and chlorfenapyr-pyrethroid LLINs compared with pyrethroid-only LLINs for malaria control in Benin: a cluster-randomised, superiority trial. Lancet. 2023;401(10375):435–46.

14. WHO. Guidelines for laboratory and field-testing of long-lasting insecticidal nets. World Health Organization; 2013. Contract No.: WHO/HTM/NTD/WHOPES/2013.3.

15. Mosha JF, Matowo NS, Kulkarni MA, Messenger LA, Lukole E, Mallya E, et al. Effectiveness of long-lasting insecticidal nets with pyriproxyfen–pyrethroid, chlorfenapyr–pyrethroid, or piperonyl butoxide–pyrethroid versus pyrethroid only against malaria in Tanzania: final-year results of a four-arm, single-blind, cluster-randomised trial. The Lancet Infectious Diseases. 2023.

16. Martin JL, Messenger LA, Mosha FW, Lukole E, Mosha JF, Kulkarni M, et al. Durability of three types of dual active ingredient long-lasting insecticidal net compared to a pyrethroid-only LLIN in Tanzania: methodology for a prospective cohort study nested in a cluster randomized controlled trial. Malar J. 2022;21(1):96.

17. WHO. Estimating functional survival of long-lasting insecticidal nets from field data. World Health Organisation. 2013.

18. Malima RC, Oxborough RMP, Tungu K, Maxwell C, Lyimo I, Mwingira V, et al. Behavioural and insecticidal effects oforganophosphate-, carbamate- and pyrethroid-treatedmosquito nets against African malaria vectors. Medical and Veterinary Entomology. 2009;23, 317–325.

19. Gillies MTC, M. A supplement to the Anophelinae of Africa south of the Sahara (Afrotropical region) 1987.

20. Koama B, Namountougou M, Sanou R, Ndo S, Ouattara A, Dabire RK, et al. The sterilizing effect of pyriproxyfen on the malaria vector Anopheles gambiae: physiological impact on ovaries development. Malar J. 2015;14:101.

21. Christophers S. The development of the egg follicle in anophelines. Paludism. 1911;2:73–8.

22. Bass C, Williamson MS, Field LM. Development of a multiplex real-time PCR assay for identification of members of the *Anopheles gambiae* species complex. Acta Trop. 2008;107(1):50–3.

23. Vezenegho SB, Bass C, Puinean M, Williamson MS, Field LM, Coetzee M, et al. Development of multiplex real-time PCR assays for identification of members of the Anopheles funestus species group. Malar J. 2009;8:282.

24. Mavridis K, Wipf N, Medves S, Erquiaga I, Muller P, Vontas J. Rapid multiplex gene expression assays for monitoring metabolic resistance in the major malaria vector Anopheles gambiae. Parasit Vectors. 2019;12(1):9.

25. Muller P, Warr E, Stevenson BJ, Pignatelli PM, Morgan JC, Steven A, et al. Field-caught permethrin-resistant Anopheles gambiae overexpress CYP6P3, a P450 that metabolises pyrethroids. PLoS Genet. 2008;4(11):e1000286.

26. Messenger LA, Matowo NS, Cross CL, Jumanne M, Portwood NM, Martin J, et al. Effects of next-generation, dual-active-ingredient, long-lasting insecticidal net deployment on insecticide resistance in malaria vectors in Tanzania: an analysis of a 3-year, cluster-randomised controlled trial. The Lancet Planetary Health. 2023;7(8):e673–e83.

27. Tungu PK, Michael E, Sudi W, Kisinza WW, Rowland M. Efficacy of interceptor® G2, a long-lasting insecticide mixture net treated with chlorfenapyr and alpha-cypermethrin against Anopheles funestus: experimental hut trials in north-eastern Tanzania. Malaria Journal. 2021;20(1):180.

28. Camara S, Ahoua Alou LP, Koffi AA, Clegban YCM, Kabran JP, Koffi FM, et al. Efficacy of Interceptor(®) G2, a new long-lasting insecticidal net against wild pyrethroid-resistant Anopheles gambiae s.s. from Côte d’Ivoire: a semi-field trial. Parasite. 2018;25:42.

29. Tchouakui M, Thiomela RF, Nchoutpouen E, Menze BD, Ndo C, Achu D, et al. High efficacy of chlorfenapyr-based net Interceptor® G2 against pyrethroid-resistant malaria vectors from Cameroon. Infectious Diseases of Poverty. 2023;12(1):81.

30. Martin JL, Mosha FW, Lukole E, Rowland M, Todd J, Charlwood JD, et al. Personal protection with PBO-pyrethroid synergist-treated nets after 2 years of household use against pyrethroid-resistant Anopheles in Tanzania. Parasites & Vectors. 2021;14(1):150.

31. Matowo NS. Differential impact of dual-active ingredient long-lasting insecticidal nets on primary malaria vectors: a secondary analysis of a 3-year, single-blind, cluster-randomised controlled trial in rural Tanzania. Lancet Planetary Health. 2023; 7 (5):E370–E80.

32. Mechan F, Katureebe A, Tuhaise V, Mugote M, Oruni A, Onyige I, et al. LLIN evaluation in Uganda project (LLINEUP): The fabric integrity, chemical content and bioefficacy of long-lasting insecticidal nets treated with and without piperonyl butoxide across two years of operational use in Uganda. Curr Res Parasitol Vector Borne Dis. 2022;2:100092.

33. Gichuki PM, Kamau L, Njagi K, Karoki S, Muigai N, Matoke-Muhia D, et al. Bioefficacy and durability of Olyset® Plus, a permethrin and piperonyl butoxide-treated insecticidal net in a 3-year long trial in Kenya. Infectious Diseases of Poverty. 2021;10(1):135.

34. Ngufor C, Fagbohoun J, Agbevo A, Ismail H, Challenger JD, Churcher TS, et al. Comparative efficacy of two pyrethroid-piperonyl butoxide nets (Olyset Plus and PermaNet 3.0) against pyrethroid resistant malaria vectors: a non-inferiority assessment. Malar J. 2022;21(1):20.

35. Soto A, Rowland M, Messenger LA, Kirby M, Mosha FW, Manjurano A, et al. Ovary Dissection Is a Sensitive Measure of Sterility in Anopheles&nbsp;gambiae Exposed to the Insect Growth Regulator Pyriproxyfen. Insects. 2023;14(6):552.

36. Toe KH, Mechan F, Tangena JA, Morris M, Solino J, Tchicaya EFS, et al. Assessing the impact of the addition of pyriproxyfen on the durability of permethrin-treated bed nets in Burkina Faso: a compound-randomized controlled trial. Malar J. 2019;18(1):383.

37. Adolfi A, Poulton B, Anthousi A, Macilwee S, Ranson H, Lycett GJ. Functional genetic validation of key genes conferring insecticide resistance in the major African malaria vector, Anopheles gambiae. Proc Natl Acad Sci U S A. 2019;116(51):25764–72.

38. Ingham VA, Wagstaff S, Ranson H. Transcriptomic meta-signatures identified in Anopheles gambiae populations reveal previously undetected insecticide resistance mechanisms. Nature Communications. 2018;9(1):5282.

39. Djouaka RF, Bakare AA, Coulibaly ON, Akogbeto MC, Ranson H, Hemingway J, et al. Expression of the cytochrome P450s, CYP6P3 and CYP6M2 are significantly elevated in multiple pyrethroid resistant populations of Anopheles gambiae s.s. from Southern Benin and Nigeria. BMC Genomics. 2008;9:538.

40. Kweyamba PA, Hofer LM, Kibondo UA, Mwanga RY, Sayi RM, Matwewe F, et al. Sub-lethal exposure to chlorfenapyr reduces the probability of developing <em>Plasmodium falciparum</em> parasites in surviving <em>Anopheles</em> mosquitoes. bioRxiv. 2023:2023.07.03.547458.

41. Sherrard-Smith E, Ngufor C, Sanou A, Guelbeogo MW, N’Guessan R, Elobolobo E, et al. Inferring the epidemiological benefit of indoor vector control interventions against malaria from mosquito data. Nat Commun. 2022;13(1):3862.

